# Health care seeking behaviour and financial protection of patients with hypertension: a cross-sectional study in rural West Bengal, India

**DOI:** 10.1101/2020.06.27.20141549

**Authors:** Sandipta Chakraborty, Rajesh Kumar Rai, Asit Kumar Biswas, Anamitra Barik, Preeti Gurung, Devarsetty Praveen

## Abstract

**Background:** Elevated blood pressure or hypertension is responsible for around 10 million annual deaths globally, and people residing in low and middle-income countries are disproportionately affected by it. India is no exception, where low rate of treatment seeking for hypertension coupled with widespread out-of-pocket payments (OOPs) have been a challenge. This study assessed the pattern of health care seeking and financial protection along with the associated factors among hypertensive individuals in a rural district of West Bengal, India.

**Method and findings:** A cross-sectional study was conducted in Birbhum district of the state of West Bengal, India during 2017-2018, where 300 individuals with hypertension were recruited randomly from a pre-defined list of individuals with hypertension in the district. Healthcare seeking along with two instance of financial protection –OOPs and relative expense, were analysed. Findings indicated that, of all hypertensives, 47% were not on treatment, 80% preferred private healthcare, and 91% of them had wide-spread OOPs. Cost of medication being a major share of expenses followed by significant transport cost to access public health care facility. Multivariable logistic regression analysis indicated longer duration of disease and private health care seeking were associated with more incident of OOPs. Results from linear regression modelling (generalized linear model) demonstrated association of co-morbidities with higher relative expenditure. Individuals belonging to poor economic group suffered from a high relative expense, compared to the richest.

**Conclusion:** This study suggested that individuals with hypertension had poor health care seeking, preferred private health care and had suboptimal financial protection. Hypertensives from economically poorer section had higher burden of health expenditure for treatment of hypertension, which indicated gaps in equitable health care for the control of hypertension.

## Introduction

Globally, non-communicable diseases (NCDs) contribute to a major share of the disease burden, where countries with differential level of development and varied phases of epidemiological transition have witnessed a significant rise in overall morbidity and mortality from NCDs [1-3]. Among all NCDs, cardiovascular diseases (ischaemic heart disease and stroke) are listed as the major cause of death worldwide, with hypertension (commonly defined as a systolic blood pressure□≥□140 or diastolic blood pressure□≥□90) being the most significant risk factor causing significant amount of premature deaths globally [4, 5]. Despite the high burden of hypertension, health system responses like health service delivery, health information and health financing for hypertension is suboptimal, especially in low and middle-income countries (LMICs) [6-10]. Evidence suggests that people seeking health care for NCDs bear significant and unjustified financial burden characterised by huge out-of-pocket payments (OOPs), often leading to irregular and absence of treatment seeking due to financial difficulties [10, 11]. In addition, studies show that overall health care seeking for blood pressure management is low and shared among public and private facilities [12, 13].

In India, between one-quarter to one-third of adults, aged 18 years or more, have hypertension. This remains a major threat to Indian healthcare system [14-16]. In the year 2010 the federal Indian government introduced the National Programme for Prevention and Control of Cancer, Diabetes, Cardiovascular Diseases and Stroke (NPCDCS) with hypertension and diabetes as the main focus areas. In addition, in 2017, the government launched the National Health Policy targeting 25% reduction in premature mortality occurring from cardiovascular diseases, cancer, diabetes or chronic respiratory diseases by 2025 [17, 18]. The main focus for research on hypertension in India is primarily on the risk factors of hypertension while few actually explored the health care utilization and service expenses among hypertensive individuals, as evidenced from the PubMed/MEDLINE database search [19-22]. From the perspective of health system strengthening and population health management, understanding the local preferences and health system capacity is essential. In this paper, we present the pattern of health care seeking, financial protection and its predictors among patients with hypertension in rural West Bengal. This study was conducted as a part of a broad study assessing the Capacity of Health Systems to combat the Emergence of Hypertension (COHESION study). In a comprehensive way COHESION study analysed blood pressure control, health care seeking, financial protection and health system responsiveness among adult hypertensive population (Unpublished data).

### Materials and Methods

#### Study setting, design and sampling

COHESION study is a population-based cross-sectional study, conducted in a population cohort of Birbhum Population Project (BIRPOP), a health and demographic surveillance system (HDSS) functioning under the ambit of Society for Health and Demographic Surveillance (www.shds.co.in), located in the Birbhum district of the state of West Bengal, India, between November 2017 and February 2018. BIRPOP spreads over four administrative blocks (namely Suri I, Sainthia, Mohammad Bazar and Rajnagar) out of a total of 19 blocks in district Birbhum. At its inception in 2008, BIRPOP included a sample of over 12,000 households selected by multistage stratified sampling method and has been periodically collecting information on indicators related to public health and demography. Till date, BIRPOP had completed three rounds of follow-up surveys, in 2008-09, 2012-13, and 2016-17 [23]. COHESION study was based on BIRPOP’s 2016-17 survey where blood pressure was measured for 12,255 individuals aged ≥ 18 years. Those recorded with high blood pressure (systolic blood pressure (SBP) ≥140 mm of Hg and/or diastolic blood pressure (DBP) ≥90 mm of Hg) or reported taking anti-hypertensive medication of any form were included in the hypertensive cohort [23, 24]. Details about the blood pressure measurement survey at BIRPOP has previously been published elsewhere [25]. Of the hypertensive cohort, 310 individuals were randomly selected for this study. Sample size was calculated using CDC Epi-info™ version 7.2, assuming 50% prevalence for hypertension control in all hypertensives, 7.5% of error and confidence interval of 99%. With the addition of 5% non-response rate, final sample size was 310 individuals of which 300 interviews were conducted. Terminally ill and mentally challenged individuals, diagnosed by a physician, were not considered for participation in the study. Data were collected by trained surveyors using Computer Assisted Personal Interview (CAPI) technique [26]. A rigorous protocol for survey monitoring was followed to assure the quality of the data being collected.

#### Outcome measurement

To understand the health care seeking behaviour, patients were asked if they were taking any medication for blood pressure control and have been visiting to any healthcare provider. Patients with a history of intake of daily medication for hypertension in the preceding four weeks were considered to be on ‘regular medication’. Those with a history of visit to any health care provider at least once in the last six months for treatment or follow-up care of hypertension, were noted to have ‘regular medical consultation’. Patients who had both of the above (regular medication and regular visit to physician) were labelled as “having regular treatment for hypertension”. Those reported only regular medication, identified as having “regular medication only”. Patients, currently not on any medication or consultation for last one year or never sought any treatment for hypertension, were labelled as “not on treatment”. The rest were categorised as ‘patients on irregular treatment”

Two outcomes in relation to cost of treatment, were analysed in this study – i) Out-of-pocket payments (OOPs), and ii) relative cost for hypertension care. Considering the varied practice of health care seeking behaviour, total expected OOPs were calculated considering multiple expenses. Expenses paid for medical consultation, transport and others like food, lodging etc. during the consultation in the last medical visit, and cost of blood pressure lowering medication if taken for a month, all together added to obtain total expected cost. Monthly per capita expenditure (MPCE) was calculated as monthly total consumer expenditure in a household over all items of consumption divided by the household size (total number of persons in the household) and was used as the proxy measure of the economic status [27]. Based on the MPCE, the participants were divided into four quartile classes and categorised into relative economic groups: poorest, lower-middle, upper-middle and richest class. Relative cost of seeking healthcare for hypertension for an individual was defined as percentage of MPCE incurred for OOPs [19].

#### Covariates

Based on existing literature from developing countries, a range of potential covariates were considered.

Social demographic characteristics: This included age in completed years (<50, 50-63, >63), gender (female and male), educational attainment (secondary and above, upper primary, primary, and illiterate/below primary), social group (other backward classes, scheduled caste/ scheduled tribes and others), religion (Hinduism and Islam), civil status (living with partner, and not living with partner), employment status (service/business, labourer, homemaker/retired/student, and unemployed), and economic status based on MPCE quartile distribution (high, upper-middle, lower-middle, and poor)

Hypertension related variables: This included duration of hypertension (<5 years, ≥5 years, and not sure/don’t know), co-morbidity (no and yes), regularity of treatment of hypertension (as elaborated before), type of health facility accessed (public and non-public), and healthcare provider like, public physician, private physician, AYUSH (*Ayurveda*, Yoga and Naturopathy, *Unani, Siddha* and Homoeopathy) doctor, and informal health care practitioner (Quack) [28]. Comorbidity refers to self-report about any of the diseases like diabetes, dyslipidaemia, chronic kidney disease or cardiovascular disease in addition to hypertension.

#### Statistical analysis

Bivariate and multivariable analyses were performed to attain the study objectives. Means and proportions were presented with 95% confidence intervals. For the purpose of regression modelling, a directed acyclic graph (DAG) was developed, based on causal diagram theory [29] and review of existing literature. Binary logistic regression was deployed to understand the predictors of OOPs, whereas linear regression by generalized linear models (GLM) was used to assess the relative costs. Measures of association were presented as odds ratio (OR) with 95% confidence intervals (CIs) with value “1” as the null point. GLM is preferred because of abundance of zero values in relative cost data and a possible non-parametric distribution [30]. With the linear modelling, the association is expressed with the estimated coefficient (Coeff) and associated 95% CIs, indicating direction of association with value “zero” as the null point. Data analysis were carried out using a statistical package - Stata, version 12.0 and *p* value was considered to interpret the significance of observed association. Qualitative interpretation based on *p* value (significant/non-significant based on conventional cut off) was judged cautiously, keeping with the study design and limitations.

#### Ethics statement

Ethical approval was granted by institutional review board of Society for health and Demographic Surveillance. Written informed consent was obtained from all participants prior to enrolment in the study. Irrespective of their participation status, all, who were approached to participate in the study were provided with a leaflet on healthy lifestyle, health education related to hypertension and other NCDs written in local language.

## Results

In total, 310 were approached to participate in this study, and 300 finally participated. **Table 1** outlines the descriptive characteristics of all the participants. The mean age of the participants was 55.99 ± 12.46 years. More than half of the participants were female and were illiterate or had not completed their primary education. Majority of the participants were Hindus and homemaker/retired/students by profession. Over 35% (n=106) of participants had hypertension for ≥5 years, and 20% (n= 60) had a co-morbid condition. Over 47% (n=141) of the participants were not on treatment, and among individuals receiving treatment, over 80% (n=128) sought healthcare from non-public healthcare provider. Over 90% (n=144) of those who sought care for blood pressure treatment incurred some OOPs. Expected cost for seeking complete care for hypertension per month was over □ 306 (> $4.5) and relative cost per month was 13.5% of the MPCE (**Table 1**). Further analysis (not shown separately) revealed that the median of relative cost was higher for those seeking care in non-public healthcare facility (median: 10.7%) compared to the public healthcare provider (2.1%). The median of OOPs was the largest for purchase of medicines (47.7%) in those seeking private healthcare, while it was transport and other costs in those seeking care (51.3%) followed by purchase of medicines (37.5%) from a public healthcare facility. (**Figure 1**).

**Table 1.** Characteristics of sampled hypertensive population.

| Background characteristics | n | mean or percentage (95% CI) |
| --- | --- | --- |
| **Age** | 300 | 55.99 (54.58-57.41) |
| **Total expected cost of seeking complete care for hypertension (₹) \*** | 159 | 306.49 (257.65-355.33) |
| **Relative cost (%) for treatment of hypertension\*** | 159 | 13.52 (11.13-15.90) |
| **Age group (years)** |  |  |
| < 50 | 101 | 33.67(28.29-39.04) |
| 50 - 63 | 107 | 35.67(30.22-41.12) |
| >63 | 92 | 30.67(25.42-35.91) |
| **Education** |  |  |
| Completed Secondary or above | 48 | 16.00(11.83-20.17) |
| Completed Upper-primary | 46 | 15.33(11.23-19.43) |
| Completed Primary | 56 | 18.67(14.23-23.10) |
| Illiterate/ Below primary | 150 | 50.00(44.31-55.69) |
| **Sex** |  |  |
| Female | 183 | 61.00(55.45-66.55) |
| Male | 117 | 39.00(33.45-44.55) |
| **Social group** |  |  |
| Others | 140 | 46.67(40.99-52.34) |
| OBC | 42 | 14.00(10.05-17.95) |
| SC/ST | 118 | 39.33(33.77-44.89) |
| **Religion<sup>+</sup>** |  |  |
| Hinduism | 225 | 75.25(70.33-80.17) |
| Islam | 74 | 24.75(19.83-29.67) |
| **Civil status** |  |  |
| Living with partner | 195 | 65.00(59.57-70.43) |
| Not living with partner | 105 | 35.00(29.57-40.43) |
| **Occupation** |  |  |
| Service/Business | 65 | 21.67(16.98-26.36) |
| Labourer | 47 | 15.67(11.53-19.80) |
| Homemaker/Retired/ Student | 160 | 53.33(47.66-59.01) |
| Unemployed | 28 | 9.33(6.02-12.64) |
| **Economic Class** |  |  |
| Richest | 75 | 25.00(20.07-29.93) |
| Upper Middle | 79 | 26.33(21.32-31.35) |
| Lower-middle | 70 | 23.33(18.52-28.15) |
| Poorest | 76 | 25.33(20.38-30.28) |
| **Duration of Hypertension (years)** |  |  |
| <5 | 141 | 47.00(41.32-52.68) |
| ≥5 | 106 | 35.33(29.89-40.77) |
| Not sure/don't know | 53 | 17.67(13.32-22.01) |
| **Co-morbidity** |  |  |
| No | 240 | 80.00(75.45-84.55) |
| Yes | 60 | 20.00(15.45-24.55) |
| **Regular treatment for hypertension** |  |  |
| On regular consultation & Medication | 71 | 23.67(18.83-28.50) |
| On regular medication only | 39 | 13.00(9.17-16.83) |
| On irregular treatment | 49 | 16.33(12.13-20.54) |
| Not on treatment | 141 | 47.00(41.32-52.68) |
| **Place of treatment for hypertension\*** |  |  |
| Public | 31 | 19.50(13.27-25.72) |
| Non-public | 128 | 80.50(74.28-86.73) |
| **Health care provider\*** |  |  |
| Public physician | 30 | 18.87(12.72-25.02) |
| Private physician | 63 | 39.62(31.94-47.31) |
| AYUSH doctor/ Other | 19 | 11.95(6.85-17.05) |
| Informal healthcare provider | 47 | 29.56(22.39-36.73) |
| **OPP\*** |  |  |
| Absent | 15 | 9.43(4.84-14.03) |
| Present | 144 | 90.57(85.97-95.16) |

₹: Indian National Rupee; CI: Confidence Interval; OBC: Other backward classes; SC: Scheduled caste; ST: Scheduled tribe; AYUSH: *Ayurveda*, Yoga and Naturopathy, *Unani*, *Siddha* and Homoeopathy; OPP: Out of Pocket Payments
\* Sample characteristics is based on 159 participants representing patients seeking treatment for hypertension
<sup>+</sup> One person did not share information on religion.

**Figure 1:**
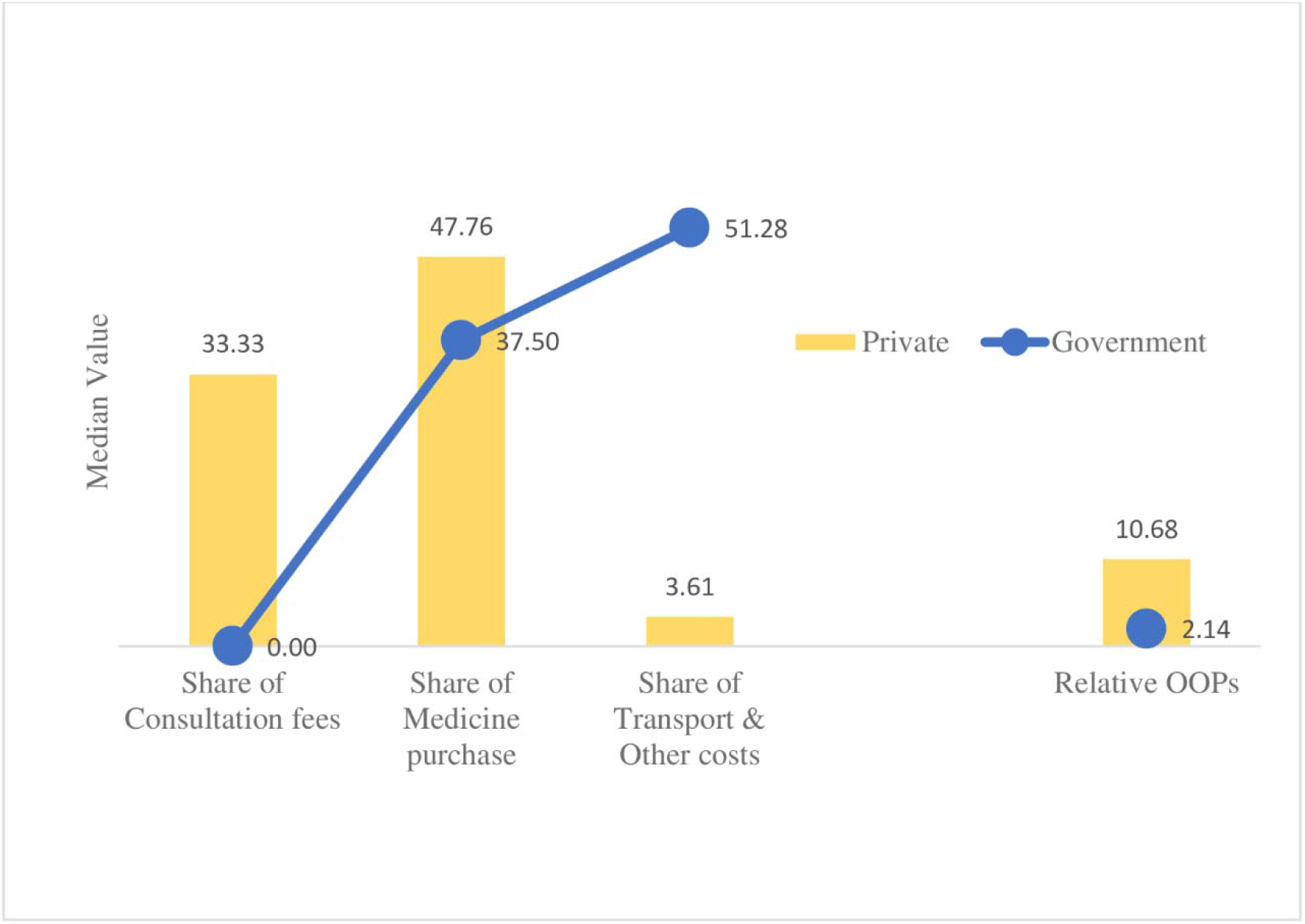
Median of OOPs share (%) and relative OOPs (%) across the place of treatment seeking

Fifteen individuals were reported incurring no OOPs for the usual treatment for hypertension. Majority (n=9) were female, aged between 50 to 63 years (n=9), Hindu (n=12), general caste (n=9) with below primary or no formal education (n=10), home maker/ retired (n=10) and belongs to upper-middle class (n=6) of the economic strata of the study population.

**Table 2** shows lower odds of having OOPs among participants aged 50-63 years and 63 years and above compared to participants below 50 years. Males when compared to females, and homemaker/retired /student, labourer and unemployed when compared to those in service/business had relatively lower odds of incurring any OOPs. Compared to the richest economic class the poorest had lower odds of having any OOPs, in unadjusted model (uOR _poorest_ 0.22 (CI: 0.04-1.21)). Having hypertension for five years or more (uOR 5.14 (CI: 1.39-19.01) and aOR 5.68 (CI: 1.24-25.99)) and seeking treatment from private establishments (uOR 26.32 (CI: 6.80-101.93) and aOR 34.33 (CI: 4.82-244.68)) were positively associated with OOPs.

**Table 2.**
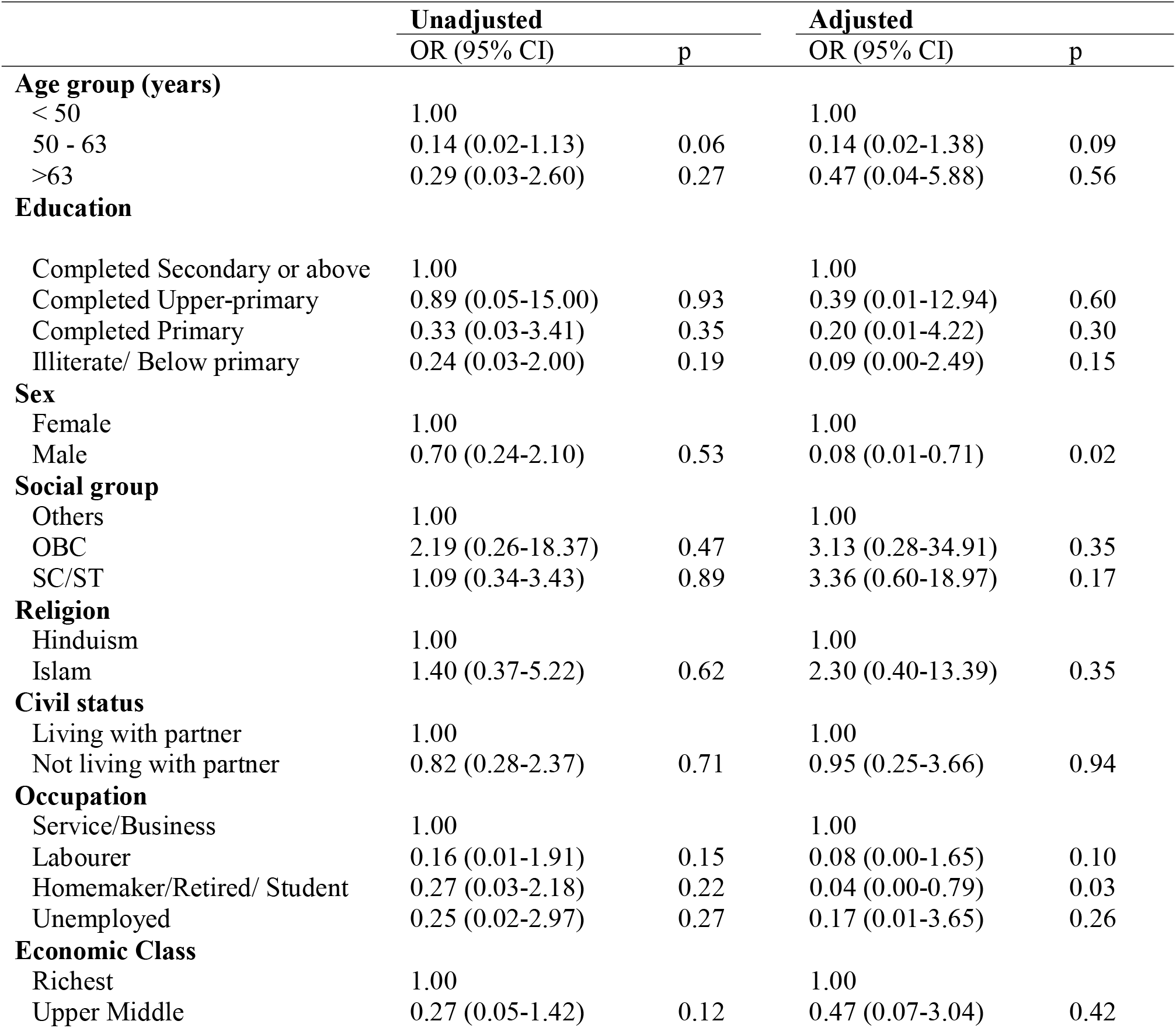
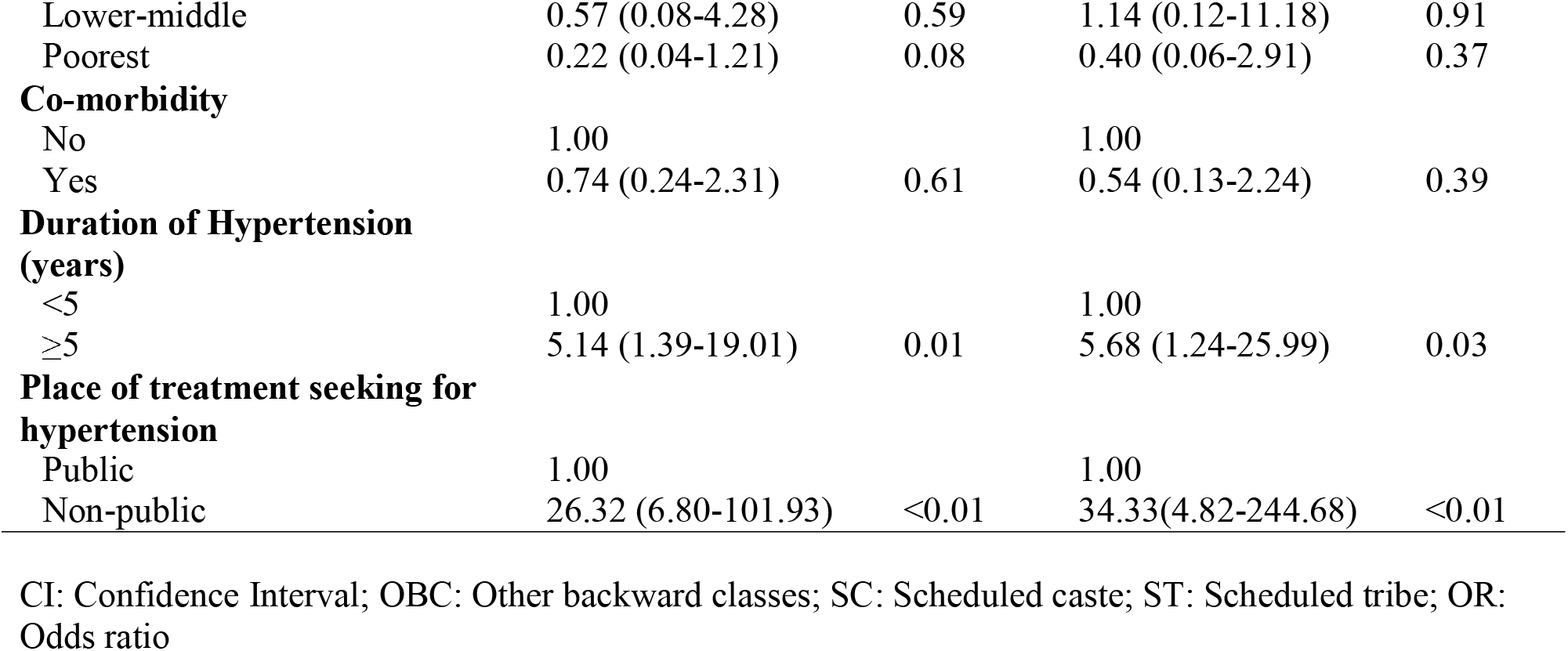
Odds of out-of-pocket payment.

Linear regression (**Table 3**) demonstrated lower relative expenses among people with primary or below level of schooling, compared to highest educational group; (Adjusted Coefficient (aCoeff) _completed primary_ −10.65 (CI: −19.78, −1.51) and aCoeff_no formal education/below primary_ −11.60 (CI: −20.88, - 2.32)). The unemployed individuals had more relative expenses compared to those engaged in service/business (Unadjusted Coefficient (uCoeff)_unemployed_ 8.71 (CI: 0.04,17.38) and aCoeff_unemployed_ 9.34 (CI: −1.74,20.43)). The poorest, lower-middle and upper-middle class had 11, 8 and 7 units of more relative expenses respectively, compared to the richest economic class (aCoeff_poorest_ 11.27 (CI: 3.82,18.71); aCoeff_lower-middle_ 7.83 (CI: 0.65,15.00) and aCoeff_upper-middle_ 7.25 (CI: 0.80,13.70)) (**Figure 2**). Presence of co-morbidity and seeking treatment from private establishments were both associated with more relative expenses (aCoeff_one or more co-morbidity_ 10.28 (CI: 4.96,15.61); reference group: no co-morbidity and aCoeff_private establishment_ 11.55 (CI: 5.74,17.37); reference group: government institution). Similarly, seeking treatment from private doctors, informal practitioners and AYUSH doctors/others were associated with more relative expenses (aCoeff_private Doctors_ 18.43 (CI: 12.13, 24.73), aCoeff_informal healthcare provider_ 5.96 (CI: −0.36, 12.28), aCoeff_AYUSH/ Other_ 10.28 (CI: 2.56, 17.99)) when compared to those seeking treatment from government doctors.

**Table 3.** Associates of relative expenses.

|  | **Unadjusted** |  | **Adjusted** |  |
| --- | --- | --- | --- | --- |
| | $\beta$ (95% CI) | p | $\beta$ (95% CI) | p |
| **Age group (years)** |  |  |  |  |
| < 50 | 0.00 |  | 0.00 |  |
| 50 - 63 | -2.39 (-8.55,3.77) | 0.45 | -3.22 (-9.84,3.40) | 0.34 |
| >63 | 0.28 (-5.78,6.33) | 0.93 | -1.66 (-8.51,5.19) | 0.64 |
| **Education** |  |  |  |  |
| Completed Secondary or above | 0.00 |  | 0.00 |  |
| Completed Upper-primary | -2.58 (-10.82,5.67) | 0.54 | -5.17 (-14.14,3.80) | 0.26 |
| Completed Primary | -3.15 (-11.02,4.73) | 0.43 | -10.65 (-19.78,-1.51) | 0.02 |
| Illiterate/ Below primary | -1.06 (-7.69,5.56) | 0.75 | -11.60 (-20.88,-2.32) | 0.01 |
| **Sex** |  |  |  |  |
| Female | 0.00 |  | 0.00 |  |
| Male | -0.94 (-5.99,4.10) | 0.71 | -3.39 (-10.90,4.13) | 0.38 |
| **Social group** |  |  |  |  |
| Others | 0.00 |  | 0.00 |  |
| OBC | 0.43 (-6.95,7.82) | 0.91 | -2.02 (-9.58,5.55) | 0.60 |
| SC/ST | 3.44 (-1.78,8.66) | 0.20 | 5.36 (-1.27,11.98) | 0.11 |
| **Religion** |  |  |  |  |
| Hinduism | 0.00 |  | 0.00 |  |
| Islam | -0.07 (-5.54,5.40) | 0.98 | 3.23 (-3.37,9.82) | 0.34 |
| **Civil status** |  |  |  |  |
| Living with partner | 0.00 |  | 0.00 |  |
| Not living with partner | 1.11(-3.68,5.90) | 0.65 | 1.85 (-3.62,7.32) | 0.51 |
| **Occupation** |  |  |  |  |
| Service/Business | 0.00 |  | 0.00 |  |
| Labourer | 5.57 (-4.40,15.54) | 0.27 | 5.88 (-5.04,16.79) | 0.29 |
| Homemaker/Retired/ Student | 1.59 (-4.38,7.56) | 0.60 | 0.85 (-7.88,9.59) | 0.85 |
| Unemployed | 8.71 (0.04,17.38) | 0.05 | 9.34 (-1.74,20.43) | 0.09 |
| **Economic Class** |  |  |  |  |
| Richest | 0.00 |  | 0.00 |  |
| Upper Middle | 5.54 (-0.38,11.46) | 0.07 | 7.25 (0.80,13.70) | 0.03 |
| Lower-middle | 5.73 (-0.97,12.43) | 0.09 | 7.83 (0.65,15.00) | 0.03 |
| Poorest | 10.39 (3.82,16.95) | 0.00 | 11.27 (3.82,18.71) | 0.00 |
| **Co-morbidity** |  |  |  |  |
| No | 0.00 |  | 0.00 |  |
| Yes | 7.74 (2.59,12.89) | 0.00 | 10.28 (4.96,15.61) | <0.01 |
| **Duration of Hypertension (years)** |  |  |  |  |
| <5 | 0.00 |  | 0.00 |  |
| ≥5 | 1.64 (-3.10,6.37) | 0.50 | 2.17 (-2.62,6.97) | 0.37 |
| **Place of treatment seeking for hypertension** |  |  |  |  |
| Public | 0.00 |  | 0.00 |  |
| Non-public | 9.35(3.56,15.14) | 0.00 | 11.55 (5.74,17.37) | <0.01 |
| **Health care provider** |  |  |  |  |
| Public physician | 0.00 |  | 0.00 |  |
| Private physician | 14.38 (8.24,20.51) | <0.01 | 18.43 (12.13,24.73) | <0.01 |
| AYUSH doctor/ Other | 5.39 (-2.72,13.50) | 0.19 | 10.28 (2.56,17.99) | 0.01 |
| Informal healthcare provider | 3.40 (-3.07,9.86) | 0.30 | 5.96 (-0.36,12.28) | 0.06 |

CI: Confidence Interval; OBC: Other backward classes; SC: Scheduled caste; ST: Scheduled tribe; AYUSH: *Ayurveda*, Yoga and Naturopathy, *Unani*, *Siddha* and Homoeopathy; OPP: Out of Pocket Payments; $\beta$ : Coefficient

**Figure 2:**
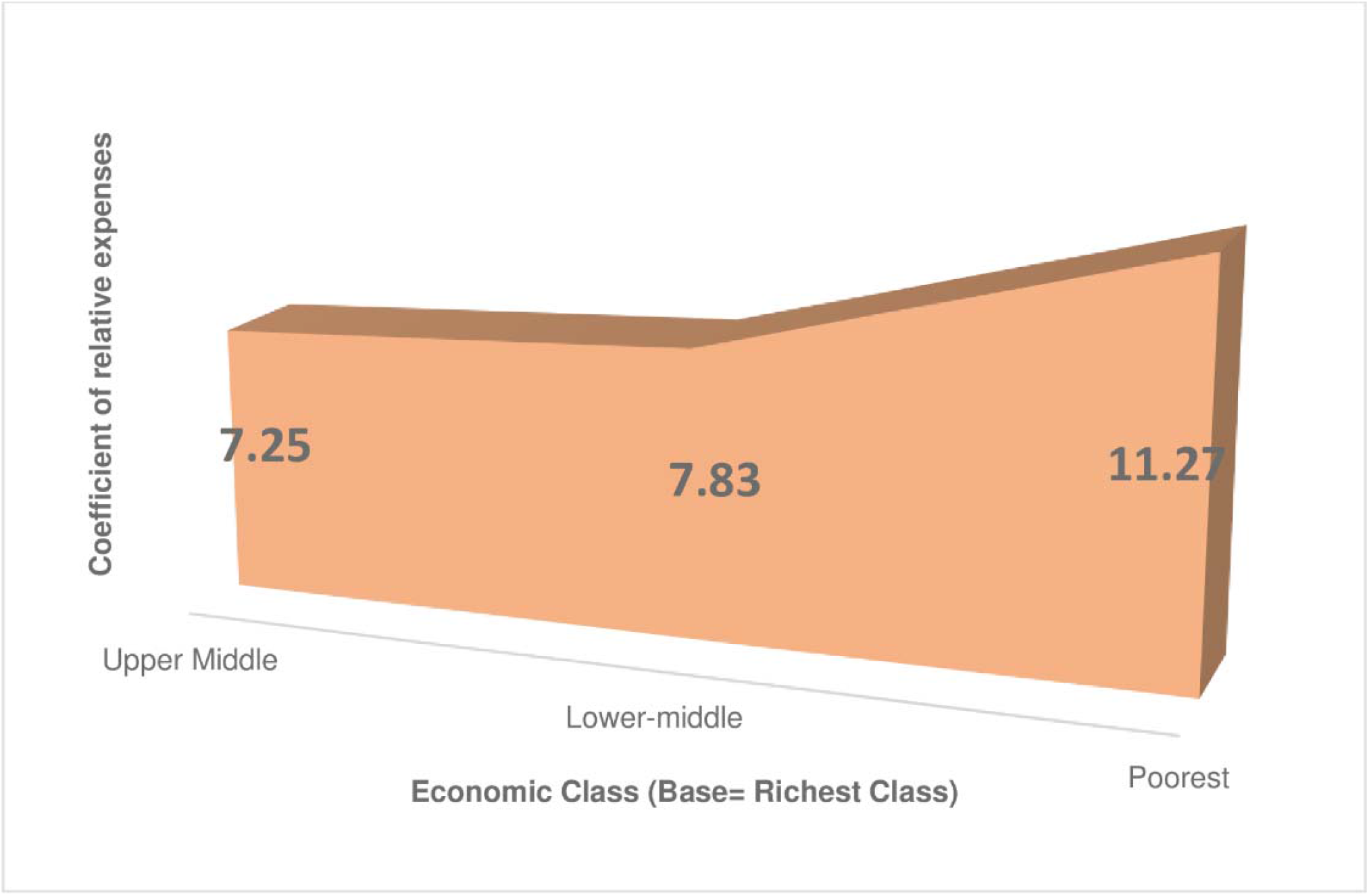
Relative expenses for care seeking across the economic strata with reference to high income group

## Discussion

India has witnessed an increasing burden of hypertension, which demands urgent attention from the public health researchers, program and policy makers. To add on to the existing body of literature on prevention of hypertension in India, this study aims to understand characteristics of healthcare seeking and financial protection among hypertensive population in West Bengal, India. The state of West Bengal recorded nearly 25% of total annual deaths and 13% of disability adjusted life years (DALYs) attributed to hypertension [15, 31]. This study revealed poor health care seeking behaviour, preference of private health facilities and high OOPs among patients who sought care for hypertension. Regression analysis adjusted for potential covariates indicate that OOPs are associated with age, sex, occupation, duration of hypertension, and place of treatment seeking for hypertension, while relative expense is associated with education, occupation, economic class, comorbidity, place of treatment and healthcare provider.

The population under study were relatively older, female predominated, had low education level, and majority were retired/homemaker. This distribution was similar to other studies where hypertension prevalence was more among elderly, females, and in poor socio-economic strata [32, 33]. The findings of poor health care seeking for blood pressure control, attributed to low awareness, affordability and availability of health care services (unpublished data). Among the hypertensives seeking treatment, OOPs were extensively reported. This scenario corroborates with previous findings of sub-optimal health system response for blood pressure control care [10, 11, 13, 14, 20, 34-37]. However better system response was associated with substantial improvement in indicators like awareness, treatment and control of hypertension in a few developed countries [38]. Similar to other studies, private establishments were major places for seeking treatment and Government institutions played a minor role for management of hypertension. Similarly, majority sought consultation from private physicians and informal healthcare providers [13, 20, 22]. The prevalence of OOPs and extent of relative cost varied between service utilization from government to non-government sources as well as with different service providers. The findings related to OOPs in this study are in line with previous reports including a report of the WHO Study on global AGEing and adult health (SAGE) but the significant variation observed in OOPs across government and private institutions in this study is found to be novel [19, 20]. Earlier studies found medicine purchase as the major share for OOPs [19-21] which corroborates with the findings from this study, however transport and other costs are also found to impose a substantial share of OOPs in Government set-up, possibly indicating better accessibility for the private treatment sources in local level compared to Government sources. This could also justify the increased usage of private facilities for hypertension management. Contrasting with findings from other studies, the present study reported lower incident of OOPs among male and those belonging to 50 years or above age group[10, 20]. Though relative cost (%) and high OOPs was proportional with level of education, the relative cost was found to be inversely related with disadvantageous economic class. These findings point towards potential issues of social justice and inequity which share a complex interrelationship [19, 20]. This might be related to poor treatment seeking behaviour among patients with low education and economic status (jointly the lower socio-economic class) owing to low awareness, financial constraint and limited access to healthcare, which may have led to lower possibility of having OOPs. But despite these barriers, patients who sought treatment experienced inequitable financial burden. Similar explanation may be applied for the unemployed group, having more extent of relative expenses while seeking care but less OOPs. Lower OOPs among homemaker/retired individuals may be due to more utilization of government health facilities, compared to the service holders/businessmen who generally have less opportunity to visit government outpatient services due to its fixed schedule. Longer duration of hypertension and existence of comorbid conditions require more intense therapy resulting in more possibility of OOPs and more relative cost (%) [10].

Limitation of the study should be interpreted in light of the results. Firstly, being a cross-sectional study, temporal ambiguity cannot be ruled out. Secondly, as most of variables under study are information based on recall, some chances for recall errors may be present. Thirdly, measurement of exact expenditure and assessing economic status could be debated. To counter the variability of health care seeking, health care expenditure related to hypertension management was calculated as expected cost for having complete care. This may have over-represented the relative cost (%) for treatment to some extent. Effects of residual confounding also cannot be ruled out. Within purview of limitations, considering the geographic and demographic uniqueness of the Birbhum population, the findings of this study should be interpreted cautiously for other settings. Despite these limitations, the study contributes tremendously to the existing literature in terms of unique study setting and use of pre-tested and validated study tools. The findings from the study suggest suboptimal financial protection of population for hypertension care. The aspect of awareness generation and evaluation of existing programs on NCDs might be needed for a better financial protection mechanism to people with hypertension.

## Data Availability

Data is available from 10.5281/zenodo.3911116

https://zenodo.org/record/3911117#.XvdkzudS8aE

## Abbreviations

aOR: adjusted odds ratio
BIRPOP: Birbhum Population Project
CI: confidence interval
NCDs: non-communicable diseases
OOPs: out of pocket payments
OR: odds ratio
uOR: unadjusted odds ratio

## Data statement

Data of COHESION study and associated codebook can be accessed through 10.5281/zenodo.3911116.

## Acknowledgements

Authors are indebted to the members of Society for Health and Demographic Surveillance, West Bengal, India, for helping execute the study.

## References

1. Abubakar I, Tillmann T, Banerjee A. Global, regional, and national age-sex specific allcause and cause-specific mortality for 240 causes of death, 1990-2013: a systematic analysis for the Global Burden of Disease Study 2013. Lancet. 2015;385(9963):117–71.

2. Habib SH, Saha S. Burden of non-communicable disease: global overview. Diabetes & Metabolic Syndrome: Clinical Research & Reviews. 2010;4(1):41–7.

3. Murray CJ, Ezzati M, Flaxman AD, Lim S, Lozano R, Michaud C, et al. GBD 2010: design, definitions, and metrics. Lancet. 2012;380(9859):2063–6.

4. World Health Organization. The top 10 causes of death 2020 [updated 9 December 2020; cited 2020, 20th December]. [Available from: https://www.who.int/news-room/fact-sheets/detail/the-top-10-causes-of-death].

5. World Health Organization. Hypertension 2019 [updated 13 September 2019; cited 2020, 20th December]. [Available from: https://www.who.int/news-room/fact-sheets/detail/hypertension].

6. Alshamsan R, Lee JT, Rana S, Areabi H, Millett C. Comparative health system performance in six middle-income countries: cross-sectional analysis using World Health Organization study of global ageing and health. Journal of the Royal Society of Medicine. 2017;110(9):365–75.

7. Feng XL, Pang M, Beard J. Health system strengthening and hypertension awareness, treatment and control: data from the China Health and Retirement Longitudinal Study. Bull World Health Organ. 2014;92(1):29–41.

8. Ibrahim MM, Damasceno A. Hypertension in developing countries. Lancet. 2012;380(9841):611–9.

9. Peck R, Mghamba J, Vanobberghen F, Kavishe B, Rugarabamu V, Smeeth L, et al.Preparedness of Tanzanian health facilities for outpatient primary care of hypertension and diabetes: a cross-sectional survey. Lancet Glob Health. 2014;2(5):e285–92.

10. Wang Q, Fu AZ, Brenner S, Kalmus O, Banda HT, De Allegri M. Out-of-pocket expenditure on chronic non-communicable diseases in sub-Saharan Africa: the case of rural Malawi. PLoS One. 2015;10(1):e0116897.

11. World Health Organization. Impact of out-of-pocket payments for treatment of non-communicable diseases in developing countries: a review of literature. 2011.

12. Bovet P, Gervasoni JP, Mkamba M, Balampama M, Lengeler C, Paccaud F. Low utilization of health care services following screening for hypertension in Dar es Salaam (Tanzania): a prospective population-based study. BMC Public Health. 2008;8(1):407.

13. Baliga SS, Gopakumaran PS, Katti SM, Mallapur MD. Treatment seeking behavior and health care expenditure incurred for hypertension among elderly in urban slums of Belgaum City. Community Med. 2013;4(2):227–30.

14. Anchala R, Kannuri NK, Pant H, Khan H, Franco OH, Di Angelantonio E, et al. Hypertension in India: a systematic review and meta-analysis of prevalence, awareness, and control of hypertension. J Hypertens. 2014;32(6):1170–7.

15. Indian Council of Medical Research; Public Health Foundation of India and Institute for Health Metrics and Evaluation. India: Health of Nation’s States - The India State-level Disease Burden Initiative. New Delhi, India: ICMR, PHFI and IHME; 2017. 2017.

16. Gupta R, Gaur K, Cv SR. Emerging trends in hypertension epidemiology in India. J Hum Hypertens. 2019;33(8):575–87.

17. Directorate General of Health Services; Ministry of Health & Family Welfare Government of India.National Programme for Prevention and Control of Cancer, Diabetes Cardiovascular Diseases and Stroke (NPCDCS) 2017 [updated 26 August 2020; cited 2021 February 22]. [Available from: http://dghs.gov.in/content/1363_3_NationalProgrammePreventionControl.aspx].

18. Ministry of Health & Family Welfare GoI. National Health Policy 2017 [cited 2021 February 22]. [Available from: https://www.nhp.gov.in/nhpfiles/national_health_policy_2017.pdf].

19. Bhojani U, Thriveni B, Devadasan R, Munegowda C, Devadasan N, Kolsteren P, et al. Out-of-pocket healthcare payments on chronic conditions impoverish urban poor in Bangalore, India. BMC Public Health. 2012;12(1):990.

20. Brinda EM, Kowal P, Attermann J, Enemark U. Health service use, out-of-pocket payments and catastrophic health expenditure among older people in India: The WHO Study on global AGEing and adult health (SAGE). J Epidemiol Community Health. 2015;69(5):489–94.

21. Engelgau MM, Karan A, Mahal A. The Economic impact of Non-communicable Diseases on households in India. Global Health. 2012;8(1):9.

22. Kanungo S, Mahapatra T, Bhowmik K, Saha J, Mahapatra S, Pal D, et al. Patterns and predictors of undiagnosed and uncontrolled hypertension: observations from a poor-resource setting. J Hum Hypertens. 2017;31(1):56–65.

23. Ghosh S, Barik A, Majumder S, Gorain A, Mukherjee S, Mazumdar S, et al. Health & Demographic Surveillance System Profile: The Birbhum population project (Birbhum HDSS). Int J Epidemiol. 2015;44(1):98–107.

24. Chobanian AV, Bakris GL, Black HR, Cushman WC, Green LA, Izzo JL, Jr., et al. The Seventh Report of the Joint National Committee on Prevention, Detection, Evaluation, and Treatment of High Blood Pressure: the JNC 7 report. JAMA. 2003;289(19):2560–72.

25. Ghosh S, Mukhopadhyay S, Barik A. Sex differences in the risk profile of hypertension: a cross-sectional study. BMJ Open. 2016;6(7):e010085.

26. Baker RPJSSCR. New technology in survey research: Computer-assisted personal interviewing (CAPI). 1992;10(2):145–57.

27. Howe LD, Galobardes B, Matijasevich A, Gordon D, Johnston D, Onwujekwe O, et al. Measuring socio-economic position for epidemiological studies in low- and middle-income countries: a methods of measurement in epidemiology paper. Int J Epidemiol. 2012;41(3):871–86.

28. Das J, Chowdhury A, Hussam R, Banerjee AV. The impact of training informal health care providers in India: A randomized controlled trial. Science (New York, NY). 2016;354(6308).

29. Rothman KJ, Greenland S, Lash TL. Modern epidemiology. 3rd ed: Lippincott Williams & Wilkins; 2008. p. 183–209.

30. Matsaganis M, Mitrakos T, Tsakloglou P. Modelling health expenditure at the household level in Greece. The European Journal of Health Economics. 2009;10(3):329–36.

31. Institute for Health Metrics and Evaluation (IHME). GBD India Compare | Viz Hub 2019 [cited 2021 February 22]. [Available from: https://vizhub.healthdata.org/gbd-compare/india].

32. Irazola VE, Gutierrez L, Bloomfield GS, Carrillo-Larco RM, Dorairaj P, Gaziano T, et al. Hypertension Prevalence, Awareness, Treatment, and Control in Selected Communities of Nine Low-and Middle Income Countries: Results From the NHLBI/UHG Network of Centers of Excellence for Chronic Diseases. Global heart. 2016;11(1):47.

33. Sarki AM, Nduka CU, Stranges S, Kandala NB, Uthman OA. Prevalence of Hypertension in Low- and Middle-Income Countries: A Systematic Review and Meta-Analysis. Medicine (Baltimore). 2015;94(50):e1959.

34. Chow CK, Teo KK, Rangarajan S, Islam S, Gupta R, Avezum A, et al. Prevalence, awareness, treatment, and control of hypertension in rural and urban communities in high-, middle-, and low-income countries. JAMA. 2013;310(9):959–68.

35. Kaur P, Rao SR, Radhakrishnan E, Rajasekar D, Gupte MD. Prevalence, awareness, treatment, control and risk factors for hypertension in a rural population in South India. Int J Public Health. 2012;57(1):87–94.

36. Singh AK, Kalaivani M, Krishnan A, Aggarwal P, Gupta SK. Prevalence, awareness, treatment and control of hypertension among elderly persons in an urban slum of Delhi, India. Indian Journal of Medical Specialities. 2014;5(1):7–10.

37. Tocci G, Ferrucci A, Pontremoli R, Ferri C, Rosei E, Morganti A, et al. Blood pressure levels and control in Italy: comprehensive analysis of clinical data from 2000–2005 and 2005– 2011 hypertension surveys. Journal of human hypertension. 2015;29(11):696.

38. Joffres M, Falaschetti E, Gillespie C, Robitaille C, Loustalot F, Poulter N, et al. Hypertension prevalence, awareness, treatment and control in national surveys from England, the USA and Canada, and correlation with stroke and ischaemic heart disease mortality: a cross-sectional study. BMJ Open. 2013;3(8):e003423.

